## Supplementary material for "Influences on policy-formulation, decision-making, organisation and management for maternal, newborn and child health in Bangladesh, Ethiopia, Malawi and Uganda: the roles and legitimacy of a multi-country network": S3_Text.docx

**QCN EVALUATION – COUNTRY CONTEXT SUPPLEMENT**

This document provides details of the context in Bangladesh, Ethiopia, Malawi and Uganda, across the whole of our QCN evaluation project and is therefore a common resource for all 9 papers in the PLOS Global Public Health QCN Evaluation collection [ref PLOSGPH collection website].

1. **BANGLADESH**

**Health system**

The health system of Bangladesh is a pluralistic system with four key actors that define the structure and function of the system: government, private sector, nongovernmental organizations (NGOs) and donor agencies [1]. The public health system is highly centralized, with planning undertaken by the Ministry of Health and Family Welfare and little authority delegated to local levels [1]. The Ministry of Health & Family Welfare (MOH&FW) is responsible for policy, planning and decision making at macro level [2]. At the central level, power is always centralized towards higher-level officials [1] and officials who are closer to project execution, do not take decisions [1,3]. Government, the private sector and NGOs are engaged in service delivery, financing and employing health staff [4]. Support from development partners has always been an important source of health financing in Bangladesh, hence donors play a key role in financing and planning health programmes [5]. There are four directorates e.g. Director General of Health Services, Director General of Family Planning, Directorate of Nursing Services and Directorate of Drug Administration, under this ministry. Community Based Health Care (CBHC) is under DGHS and in the 4th Health Population and Nutrition Sector Program the dimension of CBHC has been extended further, consisting of all health facilities within an Upazila (sub-district) [3]. Health Economics unit (HEU) is the policy and advocacy unit of the government and Quality Improvement Secretariat (QIS) is housed within HEU, and played a steering role for QCN activities in support of MOHFW. The Ministry of Health and Family Welfare, through the Directorates General of Health Services (DGHS) and Family Planning (DGFP), manages a dual system of general health and family planning services through District Hospitals (secondary level), Upazila Health Complexes, Union Health and Family Welfare Centres, and community clinics (primary level) at ward level [1]. Tertiary level hospitals are specialised hospitals, at the national level and medical colleges, at the divisional level, under DGHS.

The Government of Bangladesh (GoB) has committed to achieving SDG targets and applies a sector-wide approach under the national Health, Population, and Nutrition Sector Plan (HPNSP) 2017–2022 to address the needs and gaps for accelerated progress in maternal, newborn and child health (MNCH) [6]. The current operational plan included activities to strengthen efforts to make home deliveries safe; provide *24/7* emergency obstetric and newborn care services at the Upazila level in phases; establish a functional referral system from community to facility level; and, increase access to and utilization of evidence-based priority newborn and child health services [6].

**QI History**

Bangladesh’s work on quality improvement long pre-dates the establishment of QCN. The first initiative, ‘Quality Assurance Project’, was piloted in some hospitals in 1994 [7]. Based on the experience of this piloting project, the quality assurance program was included in the Health and Population Sector Program (HPSP 1993-2003) and continued until 2010 [7]. Subsequently, the paradigm shifted to a ‘Quality Improvement Approach’ [7], with attention to quality growing with the adoption of the healthcare financing strategy (2012-2032), which served as a roadmap to achieving Universal Health Coverage (UHC). Several respondents stressed the government’s long historical commitment to quality improvement. One respondent added that Total Quality Management (TQM) was there before joining the QCN.

The government’s commitment to quality improvement is best reflected in the inception of the Quality Improvement Secretariat (QIS), established by the Ministry of Health and Family Welfare (MoH&FW) in January 2015, and housed within the Health Economics Unit (HEU) it serves as a formal management body of the National QI committee. The Secretariat supports quality improvement (QI) initiatives across the country and strengthens and coordinates QI activities in the public and private health sector. QIS was also involved in the development of a National Quality Strategy for the health sector in 2015 [7]. Its stated mission is to achieve an effective health system that provides the highest quality of care though a quality improvement approach. This marked a shift from the third national health sector plan, where “raising more service and coverage” was the key priority. Respondents’ note how the Plan has since shaped government strategy, policy, and resources — all of which are currently focused on quality improvement, and that this aligns with the efforts of the QCN. QIS also has the experience to lead the Mother Baby Friendly Facility Initiative, with the support of a BMGF-UNICEF partnership, which aimed to contribute to the reduction of maternal and newborn deaths. The project provides a unique opportunity to test the Every Mother, Every Newborn (EMEN) Standards adapted for the country context. Respondents noted that while these quality improvement efforts have been long-standing in Bangladesh, the QCN is likely to help further coordinate and ultimately amplify pre-existing quality improvement efforts.

**Emergence and Involvement of different partners**

There was some variation among respondents regarding how Bangladesh became involved with QCN. Some respondents described it as a joint effort by the government (and specifically Bangladesh’s Ministry of Health and Family Welfare) and global partners including WHO, UNICEF, and Save the Children. Others saw these partners as inviting Bangladesh to be part of the network, and the Ministry of Health accepted and committed given existing efforts and priorities around quality improvement. One respondent reflected on how Bangladesh was initiated into QCN. Still other respondents described Bangladesh’s involvement in QCN a function of joint efforts by the Government and WHO. Several respondents perceived Bangladesh as a pioneer country in quality improvement, given its early decision to become part of the network – QCN emerged in Bangladesh from 2016.

The Quality Improvement Secretariat (QIS) took the lead of QCN activities though Unicef and Save the Children carried out the main activities, playing the role of financial, technical and implementing partner. USAID financed Save the Children and also supported technically. IHI had a major contribution, being in the consortium with Save the Children. Besides the mentioned institutions, National Institute for Preventative and Social Medicine (NIPSOM), DGHS and DGFPA, district-level Civil Surgeons were also involved in network activities. UNFPA was involved in MPDSR related activities at the national level. Some local partners e.g. Simantik worked in implementation along with Save the Children.

2. **ETHIOPIA**

**Health System**

Ethiopia has a decentralized [8] health system across 11 regional states and two city administrations [9,10]. The Federal Ministry of Health (FMOH) developed policy and technical guidelines, which are managed by Regional or City Administration Health Bureaus (RHB) [10]. A three tier health system is run by the ministry and region – specialised hospitals are at tertiary level, general hospitals at secondary level, and primary hospitals, health centres and health posts are at primary level [10].

In Ethiopia, the decline in maternal and neonatal mortality was modest though it’s remaining high. Consequently, MNCH has been prioritised in the second Health Sector Transformation Plan (HSTP-II) July 2020–June 2025, aligning with global commitment i.e. SDG3, and also having ambition to achieve Universal Health Coverage. Quality in health care has been included in the current HSTP-II with the major initiatives - institutionalize a concept of health care quality and practice at all levels, establish a National Health Care Quality Council, standardize and strengthen health care quality structures and their functions, reform, redefine, and standardize scopes and functions of health facilities, establish and strengthen collaborative, high-quality learning platforms, develop and implement a support package for public and private health facilities for accreditation, establish quality improvement hubs and strengthen regular quality of care measurement and improvement [9].

The Ministry of Health (MOH) and Regional Health Bureaus (RHBs) used an active participatory process to develop the HSTP-II [10]. The health sector first identified the nation’s long-term strategic goals in consultation with the National Plan and Development Commission. Using the Strategic Planning and Management tool, a series of consultations were held with the private sector, academia, professional associations, other government sectors and development partners [10]. These consultations are instrumental in developing a comprehensive plan and ensuring commitment and shared vision among all stakeholders, and the resulting feedback was incorporated in the plan [10].

**QI History**

Ethiopia was selected by the WHO because of previous MNCH quality improvement initiatives in the Medical Services Directorate (later renamed as Health Service Directorate).  The quality initiative, National Healthcare Quality Strategy (NHQS), had been launched in 2016, followed by the establishment of quality units at federal, regional, district and facility levels. Ethiopia was one of the few countries that already had an NHQS. Prior to the NHQS, in 2015, the Health Sector Transformation Plan (HTSP), which is part of the Ethiopian Growth and Transformation Plan II, was inaugurated. There are 5 focus areas in NHQS in which MCH is the first priority, followed by nutrition, clinical and surgical services.

Networking was not a new idea in Ethiopia, similar networking initiatives in the country, including the Ethiopian Hospitals Alliance for Quality (EHAQ), established by MoH in 2010, that aims to improve the effectiveness of healthcare through collaborative learning and clustering of facilities through referral, mainly where resources are limited and in large geographic areas. EHAQ was established and implemented across all regions to improve the quality of hospitals in Ethiopia. Ethiopian hospital alliance for quality is a system where a leading hospital is selected to support other facilities clustered under it. Examples of other quality improvement strategies include the agreement between the MoH and the Institute for Healthcare Improvement (IHI) signed in 2015 to focus on maternal and newborn mortality reduction in the country.

Further, ICARE (I-improved, C-compassionate care, A-access to health service, R-re-design the system, E-engage key stakeholders) and SaLTS, focus on improving quality services across the health system. ICARE as a quality improvement initiative that aims to reduce hospital waiting time and improve surgical volume, client experience, client engagement, and the efficient utilization of resources. In addition, the MoH launched Saving Lives through Safe Surgery (SaLTS) in 2017. The initiative was designed to make essential surgical and anaesthesia care accessible to all in the country.

In general, it was thought that the idea of coordinating, integrating, pooling together and complementing the different quality service improvement initiatives in the country helped the establishment of QCN in Ethiopia. That is, there were quality initiatives by MoH, IHI, WHO, and CHAI that could be brought together through the QCN initiative. Participants reported that, at its emergence and implementation, QCN considered the plan of the government priorities and aimed to be in alignment: QCN was a program run by the government focused on QED (quality, equity and dignity), and it was also a quality improvement initiative. MCH is the first priority in the government’s quality strategy. So, the work of this network is one of the government’s agenda. The uniqueness of this network is its focus on MCH, supported by partners and sharing experience with other facilities. It is part and parcel of the government’s strategy on quality.

**Emergence and Involvement of different partners**

In 2017, the Ministry of Health in collaboration with WHO had further discussion and established QCN in Ethiopia. Led by MoH, interested partners were identified, forums created, learning sites selected and resources mobilized. By recognizing the idea MoH established a technical working group (TWG). Then TWG prepared a national roadmap called LALI (L-leadership, A-accountability, L-learning, I-implementation) and identified learning facilities.

WHO was considered as the key actor playing vital roles in initiating, directing and coordinating the implementation of QCN at global, national and local/health facility level. WHO was known for its technical and financial support and considered as one of the potential owners and ally of the network.  At the national level, WHO introduced the network and played the leading role, and like other partners they have been providing technical and financial support to the learning facilities. USAID, UNFPA and UNICEF were also major financial and technical supporters at the federal level.

While some partners provided support at the federal level, there are also Subnational -level partners who aimed to put globally agreed-upon directives into action. WHO, Transform HDR and PHC, CHAI, and IHI were the commonly mentioned partners functioning at subnational level. The support varied from technical to financial based on the partner’s engagement. However, during our second round of data collection a respondent mentioned that eventually CHAI and IHI projects had been phased out.

**Political context**

Since 2020, political conflict has arisen in Ethiopia, particularly in the northern part, which had a huge impact on QCN activities [11].

3. **MALAWI**

**Health System**

Malawi has a decentralised [12] health system which is largely dependent on donor aid [13]. The health care delivery system mainly consists of government facilities which are free-of-charge at the point of use [14,15].

Malawi’s health care system is organised at three levels namely: primary, secondary and tertiary [14,15]. These different levels are linked to each other through an established referral system [14]. Primary and secondary level care falls under district councils [14]. The District Health Officer (DHO) is the head of the district health care system and reports to the District Commissioner (DC) who is the Controlling Officer of public institutions at district level [15]. The functions of the central level (MoH) include policy making, standards setting, quality assurance, strategic planning, resource mobilisation, technical support, monitoring and evaluation and international representation [14]. Five Zonal Health Support Offices (ZHSOs) are an extension of the central level and provide technical support to districts [14].

Malawi was one of the few countries to achieve its Millennium Development Goal target for child health [12]. It adopted the Every Newborn Action Plan in 2015 and is currently reviewing its Reproductive, Maternal, Neonatal, Child and Adolescent Health (RMNCAH) strategy. Institutional deliveries have increased significantly in the past few years, and have outpaced increases in skilled human (and material) resource availability. Quality of care for many mothers and newborns remains poor as a result. The Government is cognizant of this and has established the Quality Management Directorate (QMD) within the Ministry of Health in 2016. QMD aims to contribute to improve health and client satisfaction via provision of quality health services and to drive the national agenda to improve quality and equity in the health sector in Malawi, as well. QMD is involved in QCN. The QMD is responsible for promoting and coordinating quality management initiatives across all levels of the health system and has developed the national Quality Management policy and strategy which is aligned with the HSSP 2017-2022. The National Health Care Quality Management Policy and Strategy outlines seven priority areas and key interventions including human resource management, leadership and governance, clinical practice, client safety, support systems, customer care and evidence-based decision making [15].

The HSSPII development process involved a broad range of stakeholders and this was done through consultative workshops, technical working groups and visits to institutions [15]. Departments and disease control programmes at Ministry of Health headquarters, District Health Offices, Central Hospitals, regulatory bodies, other Ministries, Departments and Agencies (MDAs), the private sector, development partners and Civil Society Organizations (SCOs) were consulted [15].

**QI History**

Several participants mentioned that the QCN in Malawi is built on previous efforts from different partners to reduce maternal and newborn mortality as part of the MDGs/SDGs and all the work done on HIV in Malawi with the impact of reducing mother-to-child transmission.  Some previous experience on MNH projects led by different stakeholders (e.g. UNICEF, Maikhanda) in different districts had introduced QI tools – such as 5S, continuous QI, TQM, mentorship – before the QCN activities were implemented. However, there was no integration (and sometimes duplication) between the different projects and no national strategy guiding QI efforts. Hence, local teams were usually familiar with some QI tools when QCN activities were introduced but the QI tools would have been different depending on the facility and stakeholders involved.

MNH is a priority area for the Malawian government and the goals of the QCN align with national priorities which facilitated uptake of QCN. Malawi’s commitment to improving MNH outcomes is demonstrated in its policies and strategies. For example, Malawi is a signatory to the Sustainable Development Goals (SDGs) and committed to achieving SDG 3 which focuses on reducing the MMR to 140 per 100,000 live births by 2030. And over the last 20 years Malawi has been working towards achieving Universal Health Coverage (UHC) which is at the core of the national health agenda between 2015 and 2030. As a result, the government has implemented several reforms as part of Malawi’s Health Sector Strategic Plan II (HSSP 2017-2022) to fast-track achievements in UHC including decentralization of the health system, autonomy of Central hospitals; introduction of National Health Insurance; changes to service level agreements; civil service reforms, national registration and the establishment of the National Community Health Strategy. The HSSP 2017-2022 also aims to improve the quality of health care services provided and ensure the quality of health facilities.

**Emergence and Involvement of different actors**

Malawi joined the network in July 2016.  The WHO invited Malawi to join the network as a result of its poor MNH indicators and Malawi was eager to join the network, as MNH and the provision of quality care is a priority area on the national health agenda. Stakeholders from the Ministry of Health perceived the QCN as an opportunity for Malawi to learn about best practices and share ideas to reduce maternal mortality and improve the quality of care offered to mothers and children.

QMD and WHO were identified as the core stakeholders leading the network and guiding its implementation, however respondents emphasized that WHO was playing a supportive role and the initiative and its associated efforts are fully driven by the QMD. Other key stakeholders at the national level included Reproductive Health Directorate (RHD), UNICEF, UNFPA and GIZ. GIZ and UNICEF supported other community-based organizations (CBOs) directly to implement QoC activities e.g. Society of Medical Doctors (SMD) and MaiKhanda. Other stakeholders who also played substantial roles at the national level included JHPIEGO, CHAI, Maikhanda, PACHA, ONSE, NEST360, INPATH, and SMD. Some of the partners involved in the QCN in Malawi were already working in partnership with the MoH on MNCH issues, including implementing programs and supporting the delivery of services.

4. **UGANDA**

**Health System**

Uganda’s health system is hierarchical and decentralised in nature. At its helm is the ministry of health (MOH) which is responsible for the development of policies, strategic planning, resource mobilisation, and supervision of all health services. Under the MOH there are several autonomous institutions including national referral hospitals, super specialised hospitals, regional referral hospitals as well as several research and logistics institutions. Health services in each of the districts are run by District Health Officers (DHOs) who are supported by Assistant District Health Officers in charge of maternal and child health (ADHOs) and a range of other officers. The district health team (DHT) plans and budgets for medicines and supplies, and infrastructure, utilities like electricity and water; recruits and supervises human resources for health; and makes decisions for health service delivery across their district. Each district often has a district referral hospital. The lower rung after this level are the health centre IVs (HCIVs) which are referral facilities at the health sub-district (county) usually run by a medical officer supported by clinical officers, nurses and midwives. Those support and supervise the health centre IIIs (HCIII) which are usually run by a midwife/nurse or clinical officer and it’s from this level upwards that maternal services are provided. These are followed by health centre IIs (HCIIs) and village health teams (VHTs) which are supported and supervised by HCIIIs to provide services at the community level. Each level is expected to refer any complex cases to the next level up.

While the rate of facility deliveries, skilled deliveries and utilization of health facilities for curative care has increased in recent years, there are gaps in the quality of care provided in Uganda. Therefore, Quality of care is central to the global agenda of ensuring health for all at all ages (SDG 3), a goal shared by the government of Uganda. The RMNCAH Sharpened Plan [16] for Uganda is the national RMNCAH policy which sets out to address existing bottlenecks to reduce MNCH morbidity and mortality. By focusing on five strategic shifts the plan concentrated on scaling up evidence-based, high impact solutions within high burden populations through a multi-sectoral approach and mutual accountability. However, its initial iteration did not place much emphasis on quality. The revised policy started in 2021 and is intended to have an increased influence on the quality of care. Providing the access to quality services is also an agenda in the last National Health Policy as well as in the Health Sector Development Plan (HSDP) 2015/16–2019/20 [17]. The development of this HSDP started with the development of the Health Issues Paper which formed the health sector contribution to the NDP II [18]. This was followed by development of the HSDP concept note which was presented to MoH Senior Management Committee and the Health Policy and Advisory Committee (HPAC) for approval in December 2014. A Taskforce was set up, chaired by the Director General of Health Services (DGHS), and having representation from all MoH departments, the 14 sector technical working groups (TWGs), Local Governments (LGs), Civil society and Non-Governmental Organizations (NGOs), the Private Sector, other Government Ministries, Departments and Agencies (MDAs), Academia, and Health Development Partners (HDPs).

**QI History**

Respondents repeatedly suggested that the QCN in Uganda built upon a long history of QI initiatives that remain ongoing, particularly those focused on HIV and reproductive health care, including ART provision and PMTCT. Participants understood this history as the reason Uganda was selected to join the QCN, as the country had QI structures in place through the Department of Quality Assurance (later, renamed to Standards Compliance Accreditation Patient Protection or, SCAPP) within the Uganda Ministry of Health and QI teams at the district and sub-district levels. At the district level, Uganda has District League Tables ranking the performance of all districts on health-related indicators, of which 50% are related to maternal health.  The adoption of various components of quality in healthcare dates back to 1994 in Uganda; it was the first country to implement quality assurance on a large scale in Africa.

Previous quality improvement (QI) initiatives in Uganda were the Yellow Star and using the 5S’s (sort, set, shine, standardize, and sustain), approach in HIV (including PMTCT), TB and malaria, which established QI teams at each level of the health system as well as specific standards, indicators and databases. More recently (2021-22) Uganda has leveraged its HIV-PMTCT history which had a nationwide database capturing data from all health facilities to include MNCH standards and indicators specific to HIV. Uganda had also started MPDSRs several years prior to the QCN starting and this aspect has continued to grow from strength to strength although while maternal death audits are more frequently undertaken and fairly seamless, perinatal death audits continue to be a major challenge at the frontline. The old structure has since changed in 2019, when quality of care work was moved under the Department of Quality Assurance, now known as SCAPP (Standards, Compliance, Accreditation, and Patient Protection). The SCAPP department worked hand in hand with the MCH and Reproductive Health (RH) departments as technical leads on MNCH issues. In line with the country’s decentralised health system there were also regional quality improvement teams (QIT) which were essentially supposed to lead on and support district QIT and health facility QITs.  However, participants reported that while this structure exists, many QI teams at the facility level were not fully operational/functional and it was not always clear the extent of ongoing activity of any of these historic QI initiatives. Regional QITs (which are comprised of DHOs within all the districts in that particular region and should meet quarterly) were also at different levels of functionality.

 	Participants found the existing structures did allow for easier implementation of the QCN, where existing general or HIV-focused QI teams could be expanded to include MNCH or latent teams could be revitalized for the QCN. Another participant at the facility level echoed this sentiment that the QCN reinvigorated existing structures.

**Emergence and Involvement of different partners**

QCN emerged in Uganda in 2017, but several noted that it was a slow start getting off the ground in the country.  Participants were in agreement that Uganda was selected to participate in the QCN due to a combination of having a history of QI efforts, having an existing structure for QI implementation embedded in the health system, and government willingness around improving quality, but also a consistently high maternal and newborn mortality rate.

At the national level, the WHO in Uganda sat between the WHO and the MoH. In addition, the WHO hired a MNCH Consultant to directly focus and provide support for QoC for MNCH – this officer, while under WHO, worked very closely with the MOH and frontline facilities. Other officers within the WHO directly supervised this consultant. WHO basically provided technical support though it also directly funded and supported some facilities and regions. USAID was involved at all levels of the network in Uganda, sitting on the technical working group at the national level. Through its Maternal Child Health and Nutrition Activity (run by FHI360) that began in 2019, USAID provided technical support to the MoH for creation and dissemination of policies, protocols, and guidelines across all areas of maternal, child health and nutrition with a particular emphasis on QI. UNICEF sits on the technical working group at the national level and oversees implementation in the northern districts. UNICEF supported implementation through its partners, including Baylor Uganda and AVSI. UNICEF operated a lot in the West Nile region where it supported several districts/facilities. For QCN in particular it was given responsibility for one of the learning sites (Kasese district) and it supported the district earlier on with activities such as training and supportive supervision. However, as the network progressed, UNICEF did not have a specific “funding pot” for Kasese which was not located in its main jurisdiction of West Nile and so support for Network activities had been marginal. UNFPA sat on the national technical working group and worked closely with the MoH MCH cluster on the QI framework and strategy. They have been involved in adapting the WHO standards and indicators for the Ugandan context and to fit within Uganda’s existing HMIS. UNFPA were also assigned to support one of the established learning districts through funding and implementation support, but UNFPA participants reported that the organisation did not have the resources to fulfil this assignment and have instead been implementing activities similar to the QCN framework in other districts where they did have resources and other ongoing activities. In Karamoja region, where UNFPA had been undertaking QI activities since 2020, rather than in Kiryandongo where they were initially assigned, UNFPA’s activities had been focused on capacity-building, including setting up QI committees at the facility and district level and training around monitoring and supervision. Over the course of 2021, UNFPA increased its involvement with QCN and decided to formally enter the network rather than mirror its work independently. As of autumn 2021, in the second round of data collection, UNFPA was finishing orientation training and making implementation plans in coordination with SCAPP and Marie Stopes.

11. Humanitarian and political situation in Ethiopia; 2022
